# Impact of sex in stroke in the young

**DOI:** 10.1101/2022.09.05.22279628

**Authors:** Anina Schwarzwald, Urs Fischer, David Seiffge, Morin Beyeler, Adrian Scutelnic, Johannes Kaesmacher, Pasquale Mordasini, Tomas Dobrocky, Jan Gralla, Mirjam R Heldner, Roza Umarova, Thomas R Meinel, Marcel Arnold, Simon Jung, Barbara Goeggel Simonetti

**Author notes:** **Corresponding author** Anina Schwarzwald, Department of Neurology, Inselspital, Bern University Hospital, and University of Bern, Freiburgstrasse, 3010 Bern, Switzerland. contributed equally (Simon Jung, Barbara Goeggel Simonetti).

## Abstract

**Background and Purpose:** Limited data is available on sex differences in young stroke patients describing discrepant findings. This study aims to investigate the sex differences in young stroke patients.

**Methods:** Prospective cohort study comparing risk factors, etiology, stroke localization, severity on admission, management and outcome in patients aged 16-55 years with acute ischemic stroke consecutively included in the Bernese stroke database between 01/2015 to 12/2018 with subgroup analyses for very young (16-35y) and young patients (36-55y).

**Results:** 689 patients (39% female) were included. Stroke in women dominated in the very young (53.8%, p<0.001) and in men in the young (63.9%, p<0.001). As risk factors only sleep-disordered breathing was more predominant in men in the very young, whereas almost all risk factors were more predominant in men in patients older than 35y. The higher incidence of stroke in women in the very young may be explained by the sex specific risk factors pregnancy, puerperium, the use of oral contraceptives, and hormonal replacement therapy. Stroke severity at presentation, etiology, stroke localization, management, and outcome did not differ between women and men.

**Conclusions:** The main finding of this study is that sex specific risk factors in women may contribute to a large extent to the higher incidence of stroke in the very young in women. Almost all modifiable stroke risk factors are more prevalent in men, either in the young as well as in the very young. These findings have major implications for primary preventive strategies of stroke in young people.

## Introduction

According to the World Health Organization (WHO), stroke is the second leading cause of death worldwide. It is also one of the most common causes of disability.^1^ Young adults up to the age of 50 account for about 10% of all stroke patients and the incidence of ischemic strokes among the latter is increasing globally.^2,3^ Despite better prognosis of stroke in the young overall, stroke in young people has an important socioeconomic impact.^3,4^

Sex differences in stroke are well known with women being older at their first-ever stroke and suffering from a poorer outcome than men.^5^ However, only few studies address sex differences in the population of young stroke patients. It is unknown if risk factors unique to women, such as pregnancy and puerperium, and the use of oral contraceptives account for the higher stroke incidence among women in the very young stroke patients (<35 years) reported by some studies.^3,6,7,8^ Only few studies addressed possible sex differences of risk factors in young patients and described that arterial hypertension, smoking, or diabetes mellitus seem to be more prevalent in men, while others, such as low physical activity and high body mass index (BMI) are more prevalent in women.^7,9^ Evidence on sex differences regarding etiology, localization, and outcome in young stroke patients is scarce. As a better understanding of sex differences provides a further step towards individualized precision medicine, this study aims to determine sex differences in young stroke patients regarding severity at presentation, stroke risk factors, stroke etiology, infarct localization, acute therapy, and outcome stratified according to age groups.

## Methods

This is a prospective, single-center cohort study, with consecutive enrolment of patients, who gave their general consent for using their personal data for research purposes, with acute ischemic stroke (AIS), transient ischemic attack (TIA), retinal infarction or transient monocular blindness aged between 16 and 55 years, who presented at the Stroke Center of the University Hospital Inselspital Bern between January 2015 and December 2018. Only patients who gave consent to use their health related data for research purposes have been included. The following variables were collected prospectively: Demographic data, data on past medical history, National Institutes of Health Stroke Scale (NIHSS)^10^ on admission, information about acute treatment (antiplatelet drugs, anticoagulants, intravenous thrombolysis (IVT) with recombinant tissue plasminogen activator (rtPA) and intra-arterial treatment (IAT) (including pharmacological, such as fibrinolysis by rtPA or urokinase, and mechanical treatment), risk factors, etiology (according to the TOAST (Trial of Org 10172 Acute Stroke Treatment) classification^11^), infarct localization (divided into anterior and posterior vascular territory), outcome after three months according to a modified Rankin Scale score (mRS)^12^, and recurrent events.

The study was approved by the institutional review board, the ethics committee of Bern, and conducted in accordance with institutional guidelines.

### Definition and data acquisition of specific variables

Persistent foramen ovale (PFO) was diagnosed by transesophageal echocardiography with adequate Valsalva maneuver. Screening for sleep-disordered breathing was performed by ApneaLink™ (by ResMed), and in case of relevant findings validated by polysomnography.The etiology of stroke was classified according to the TOAST classification.^11^ If a stroke was only associated with a PFO as a possible cause, the stroke’s etiology was classified as “undetermined”.

### Statistics

The statistical analysis was performed with IBM SPSS® version 25. Fisher’s exact test or Pearson χ^2^ test were used for categorical variables and Mann-Whitney *U* test for continuous variables. We performed subgroup analysis for the 16-35 and 36-55 year age groups. A two-tailed probability value < 0.05 was considered statistically significant.

## Results

### Demographics

Between January 2015 and December 2018, 689 consecutive patients were included in this analysis: 554 AIS (80.4%), 126 TIA (18.3%), six transient monocular blindness (0.9%) and three with retinal infarction (0.4%). Men accounted for most patients overall (60.8%), while 53.8% of the very young patients were women.

### Risk factors

In our study cohort, modifiable risk factors were more prevalent in men, such as dyslipidemia (119 women (45.4%) vs. 238 men (57.9%), p-value = 0.002), smoking (87 women (33.1%) vs. 184 men (45.0%), p-value = 0.002), coronary heart disease (5 women (1.9%) vs. 27 men (6.5%), p-value = 0.005), overweight (BMI ≥ 25 kg/m^2^; 57 women (21.4%) vs. 167 men (40.3%), p-value < 0.001), and sleep-disordered breathing (18 women (10.2%) vs. 71 men (26.5%), p-value < 0.001).

Family history of stroke and/or coronary heart disease did not differ (table 1A). Although no sex difference was found for persistent foramen ovale, women had more often a Risk of Paradoxical Embolism (RoPE)-score^13^ > 7 than men (23 of 61 documented RoPE-Scores > 7 in women (37.7%) vs. 18 of 107 in men (16.8%), p = 0.005).

**Tab. 1 A:**
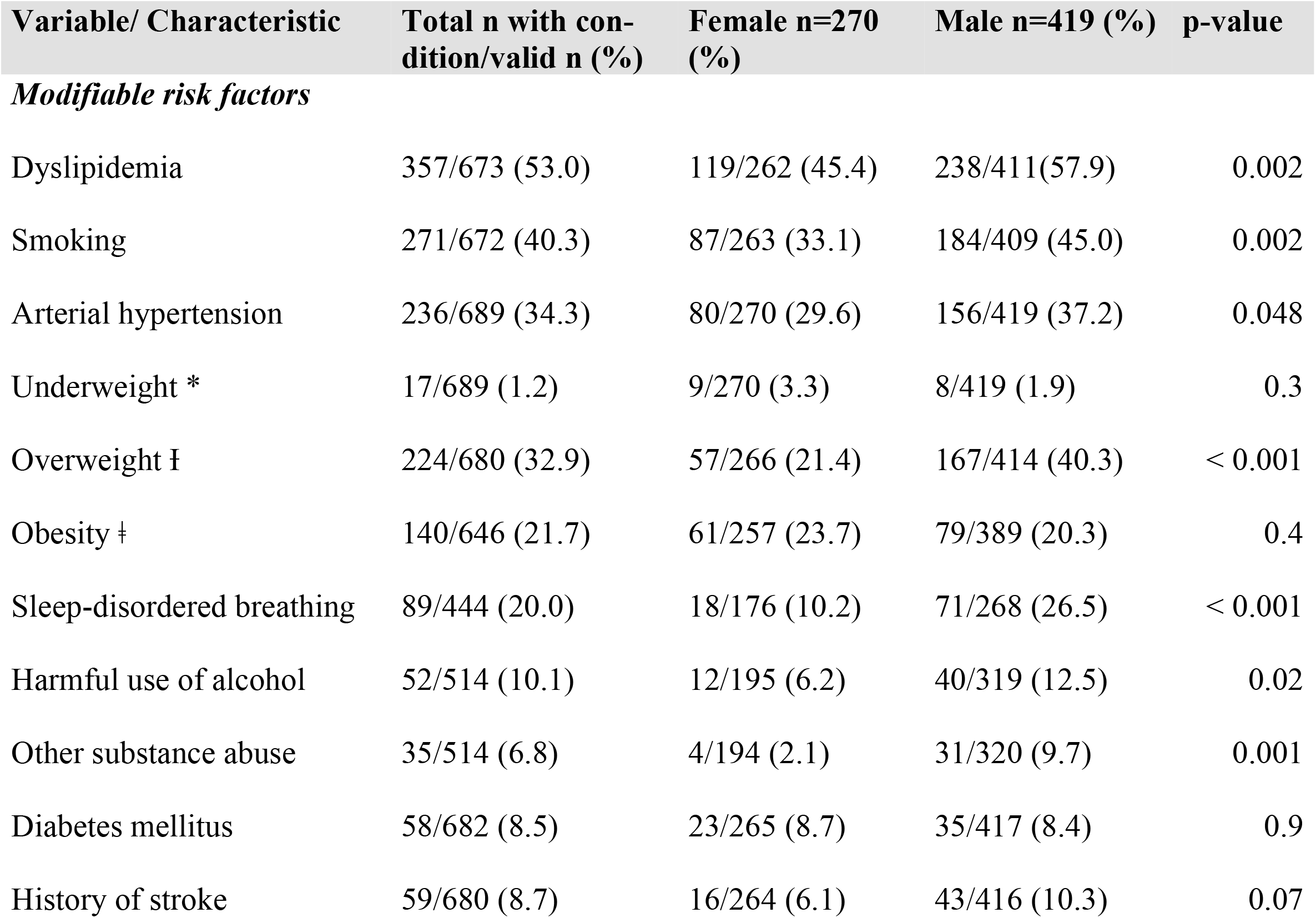
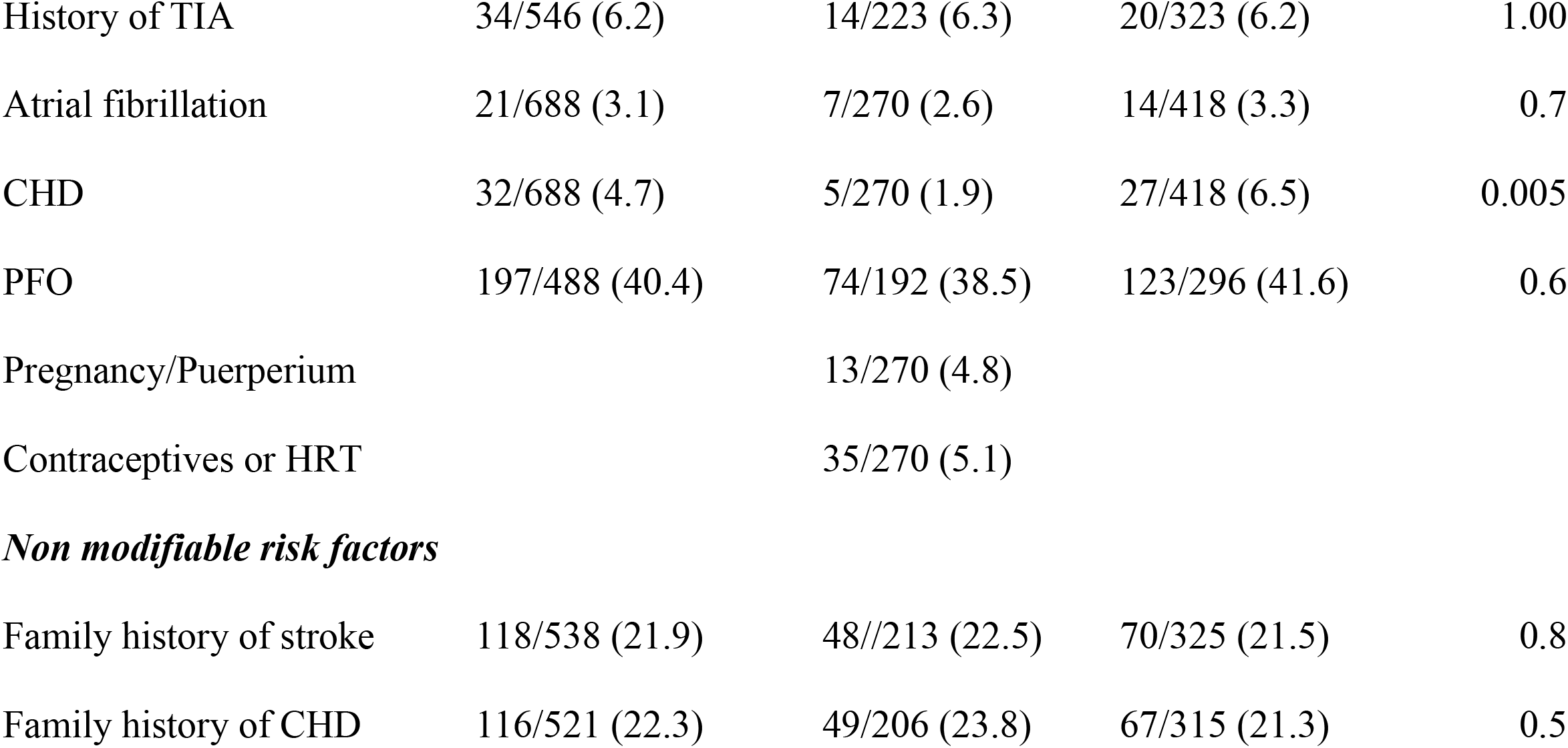
Risk factors stratified according to sex

| Variable/ Characteristic | Total n with condition/valid n (%) | Female n=270 (%) | Male n=419 (%) | p-value |
| --- | --- | --- | --- | --- |
| *Modifiable risk factors* |  |  |  |  |
| Dyslipidemia | 357/673 (53.0) | 119/262 (45.4) | 238/411(57.9) | 0.002 |
| Smoking | 271/672 (40.3) | 87/263 (33.1) | 184/409 (45.0) | 0.002 |
| Arterial hypertension | 236/689 (34.3) | 80/270 (29.6) | 156/419 (37.2) | 0.048 |
| Underweight * | 17/689 (1.2) | 9/270 (3.3) | 8/419 (1.9) | 0.3 |
| Overweight † | 224/680 (32.9) | 57/266 (21.4) | 167/414 (40.3) | < 0.001 |
| Obesity ‡ | 140/646 (21.7) | 61/257 (23.7) | 79/389 (20.3) | 0.4 |
| Sleep-disordered breathing | 89/444 (20.0) | 18/176 (10.2) | 71/268 (26.5) | < 0.001 |
| Harmful use of alcohol | 52/514 (10.1) | 12/195 (6.2) | 40/319 (12.5) | 0.02 |
| Other substance abuse | 35/514 (6.8) | 4/194 (2.1) | 31/320 (9.7) | 0.001 |
| Diabetes mellitus | 58/682 (8.5) | 23/265 (8.7) | 35/417 (8.4) | 0.9 |
| History of stroke | 59/680 (8.7) | 16/264 (6.1) | 43/416 (10.3) | 0.07 |
| History of TIA | 34/546 (6.2) | 14/223 (6.3) | 20/323 (6.2) | 1.00 |
| Atrial fibrillation | 21/688 (3.1) | 7/270 (2.6) | 14/418 (3.3) | 0.7 |
| CHD | 32/688 (4.7) | 5/270 (1.9) | 27/418 (6.5) | 0.005 |
| PFO | 197/488 (40.4) | 74/192 (38.5) | 123/296 (41.6) | 0.6 |
| Pregnancy/Puerperium |  | 13/270 (4.8) |  |  |
| Contraceptives or HRT |  | 35/270 (5.1) |  |  |
| ***Non modifiable risk factors*** |  |  |  |  |
| Family history of stroke | 118/538 (21.9) | 48//213 (22.5) | 70/325 (21.5) | 0.8 |
| Family history of CHD | 116/521 (22.3) | 49/206 (23.8) | 67/315 (21.3) | 0.5 |

The sex specific risk factors pregnancy and puerperium were found in 13/270 women (4.8%), and 35 (13%) were users of oral contraceptives or hormone replacement therapy on admission. In contrast to the analysis of our study cohort, the subgroup analysis of the 16–35-year-old patients revealed only an increased incidence of sleep-disordered breathing in men (0 women (0.0%) vs. 7 men (16.7%), p-value = 0.01) and the sex specific risk factors pregnancy and puerperium in women. In 36-55-year-old patients, most risk factors were more frequent in men (Table 1B and C).

**Tab. 1 B:** Risk factors stratified according to sex in very young AIS patients (16-35y)

| Variable/ Characteristic | Total n with condition/valid n (%) | Female (n=64) | Male (n=55) | two-tailed p-value |
| --- | --- | --- | --- | --- |
| *Modifiable risk factors* |  |  |  |  |
| Arterial hypertension | 9/119 (7.6) | 3/64 (4.7) | 6/55 (10.9) | 0.3 |
| Diabetes mellitus | 1/116 (0.9) | 0/61 (0.0) | 1/55 (1.8) | 0.5 |
| Dyslipidemia | 35/116 (30.2) | 16/62 (25.8) | 19/54 (35.2) | 0.3 |
| Smoking | 39/17 (33.3) | 17/63 (27.0) | 22/54 (40.7) | 0.1 |
| Atrial fibrillation | 0/119 (0.0) | 0/64 (0.0) | 0/55 (0.0) | 1.0 |
| CHD | 0/119 (0.0) | 0/64 (0.0) | 0/55 (0.0) | 1.0 |
| History of stroke | 10/116 (8.6) | 5/61 (8.2) | 5/55 (9.1) | 1.0 |
| History of TIA | 6/96 (6.3) | 3/52 (3.1) | 3/44 (3.1) | 1.0 |
| Underweight* | 5/113 (4.4) | 3/61 (4.9) | 2/52 (3.8) | 1.0 |
| Overweight † | 29/113 (25.7) | 13/61 (21.3) | 16/52 (30.8) | 0.3 |
| Obesity ‡ | 17/113 (15.0) | 6/61 (9.8) | 11/52 (21.2) | 0.1 |
| Sleep-disordered breathing | 7/82 (8.5) | 0/40 (0.0) | 7/42 (16.7) | 0.01 |
| Alcohol overuse | 1/89 (1.1) | 0/48 (0.0) | 1/41 (2.4) | 0.5 |
| Substance abuse | 3/89 (3.4) | 0/48 (0.0) | 3/41 (7.3) | 0.09 |
| PFO | 36/80 (45.0) | 17/39 (43.6) | 19/41 (46.3) | 0.9 |
| Pregnancy/Puerperium |  | 12/61 (19.6) |  |  |
| Contraceptives or HRT |  | 15/61 (24.6) |  |  |
| *Non modifiable risk factors* |  |  |  |  |
| Family history of stroke | 22/102 (21.6) | 14/57 (24.6) | 8/45 (17.8) | 0.5 |
| Family history of CHD | 15/100 (15.0) | 7/55 (12.7) | 8/45 (17.8) | 0.6 |

**Tab. 1 C:** Risk factors stratified according to sex in young AIS patients (36-55y)

| Variable/ Characteristic | Total n with condition/valid n (%) | Female (n=206) | Male (n=364) | two-tailed p-value |
| --- | --- | --- | --- | --- |
| ***Modifiable risk factors*** |  |  |  |  |
| Arterial hypertension | 227/570 (39.8) | 77/206 (37.4) | 150/364 (41.2) | 0.4 |
| Diabetes mellitus | 57/566 (10.1) | 23/204 (11.3) | 34/362 (9.4) | 0.5 |
| Dyslipidemia | 322/557 (57.8) | 103/200 (51.5) | 219/357 (61.3) | 0.03 |
| Smoking | 232/555 (41.8) | 70/200 (35.0) | 162/355 (45.6) | 0.02 |
| Atrial fibrillation | 21/569 (3.7) | 7/206 (3.4) | 14/363 (3.9) | 1.0 |
| CHD | 32/569 (5.6) | 5/206 (2.4) | 27/363 (7.4) | 0.01 |
| History of stroke | 49/564 (8.7) | 11/203 (5.4) | 38/361 (10.5) | 0.04 |
| History of TIA | 30/460 (6.5) | 11/169 (6.5) | 19/291 (6.5) | 1.0 |
| Underweight * | 12/527 (2.3) | 6/193 (3.1) | 6/334 (1.8) | 0.4 |
| Overweight † | 197/527 (37.4) | 44/193 (22.8) | 153/334 (45.8) | <0.001 |
| Obesity ‡ | 123/527 (23.3) | 55/193 (28.5) | 68/334 (20.4) | 0.04 |
| Sleep-disordered breathing | 82/421 (19.5) | 18/152 (11.8) | 64/269 (23.8) | 0.003 |
| Alcohol overuse | 51/425 (12.0) | 12/147 (8.2) | 39/278 (14.0) | 0.09 |
| Substance abuse | 32/425 ((7.5) | 4/146 (2.7) | 28/279 (10.0) | 0.006 |
| PFO | 161/408 (39.5) | 57/153 (37.3) | 104/255 (40.8) | 0.5 |
| Pregnancy/Puerperium |  | 1/206 (0.5) |  |  |
| Contraceptives or HRT |  | 20/206 (9.7) |  |  |
| ***Non modifiable risk factors*** |  |  |  |  |
| Family history of stroke | 96/436 (22.0) | 34/156 (21.8) | 62/280 (22.1) | 1.0 |
| Family history of CHD | 101/421 (24.0) | 42/151 (27.8) | 59/270 (21.9) | 0.2 |

TIA indicates transient ischemic attack; PFO, persistent foramen ovale; HRT, hormone replacement therapy; CHD, coronary heart disease
Body Mass Index (BMI)
\* BMI < 18.5 kg/m<sup>2</sup>
† BMI < 30 and ≥ 25 kg/m<sup>2</sup>
‡ BMI ≥ 30 kg/m<sup>2</sup>

### Stroke etiology

Stroke etiology did not differ between the sexes, neither in the whole cohort nor within the two age groups. Of note, the cause of stroke in young adults remained most often undetermined (403 (58.7%) of 687 in total, 164 of 269 women (60.9%) vs. 239 of 418 men (57.2%)). Other determined etiologies were found second most frequently (143 (20.8%), 57 women (21.2%) vs. 86 men (20.6%)), followed by strokes due to cardioembolism (70 in total (20.2%), 23 women (8.6%) vs. 47 men (11.2%)) (Figure 2).

**Fig. 1:**
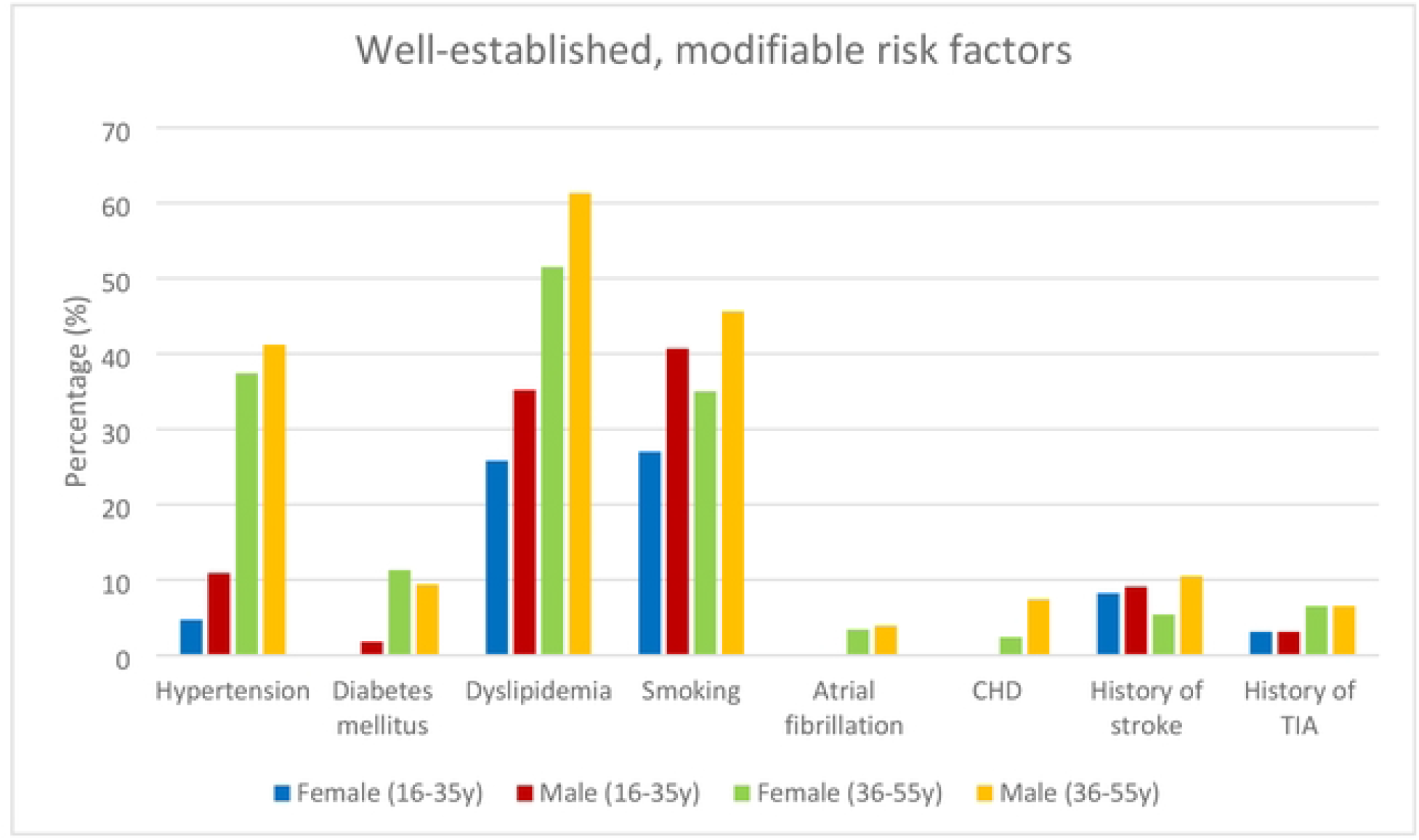
Proportion of well-established risk factors stratified according to sex and age in young stroke patients

**Fig. 2:**
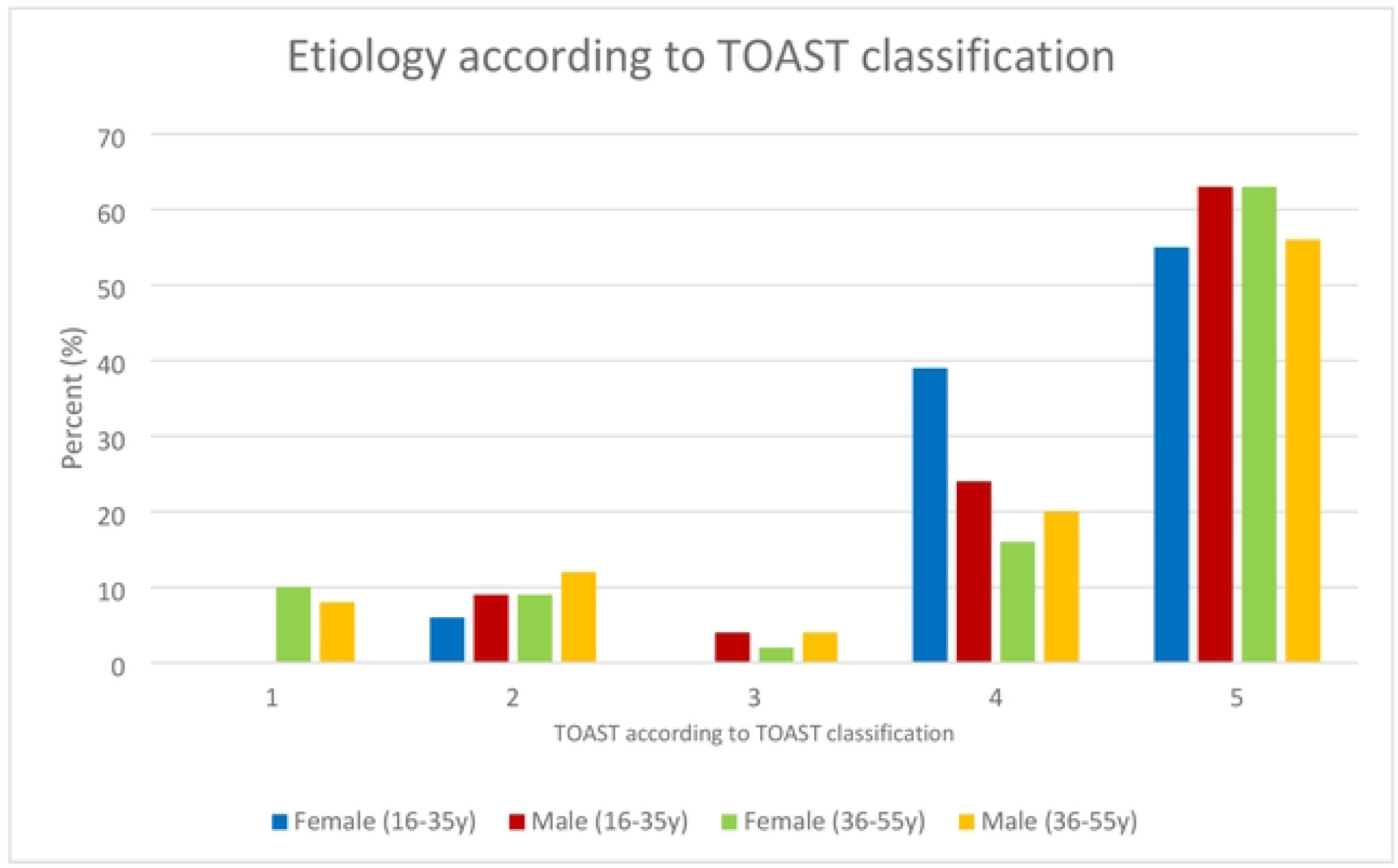
Etiology according to TOAST criteria^11^ distributed according to sex and age

### Infarct localization

Men and women did not differ in terms of stroke localization, neither in the whole study population, nor in the two age groups (175 of 270 women (65%) and 282 of 419 men (67%) with anterior circulation stroke: 95 women (35%) and 137 men (33%) with posterior circulation stroke (p=0.5)). In addition, stroke localization in the age subgroups did not differ (anterior circulation in 58% of women and 62% of men in the 16-35year age group (p=0.7); 67% of women and 68% of men in the 36-45year age group (p=0.8)).

### Stroke severity on admission and acute treatment

Stroke severity on admission did not differ between the sexes (mean NIHSS women: 4.61, men: 4.65, p=0.56), nor the choice of acute treatment (Table 2).

**Tab. 2:** Treatment procedures according to sex

|  | **Valid n/total**<br>**(t=689) (%)** | **Female (t=270)**<br>**(%)** | **Male (t=419)**<br>**(%)** | **Two-tailed**<br>**p-value** |
| --- | --- | --- | --- | --- |
| Antiplatelet drugs | 387 (56.2) | 154 (57.0) | 233 (55.6) | 0.8 |
| Anticoagulants | 48 (7.0) | 17 (6.3) | 31/419 (7.4) | 0.6 |
| IVT with rTPA | 126 (18.3) | 40 (14.8) | 86 (20.5) | 0.07 |
| IAT | 138 (20.0) | 54 (20.0) | 84 (20.0) | 1.0 |

IVT indicates intravenous thrombolysis; rTPA, recombinant tissue plasminogen activator; IAT, intra-arterial treatment

### Clinical outcome after three months

A majority of both men (87%) and women (87%) had a good outcome defined as a mRS ≤ 2 (p=0.8). Case fatality did not differ (4.0% in women and 4.6% in men, p=0.84), nor did recurrent ischemic events (4.4% in women vs. 2.9% in men, p =0.39).

## Discussion

The main finding of our study is that men have a higher incidence of sleep-disordered breathing in the 16-35 years old stroke patients, but the higher incidence of stroke in women in this age group may largely be explained by sex specific risk factors, including pregnancy, puerperium and the use of oral contraceptives. In contrast, in patients older than 35, there is a male predominance of almost all risk factors. We did not find sex differences regarding stroke etiology, severity at presentation, localization, and outcome in stroke in the young.

In accordance with previous studies, we found a female predominance in the age group 16-35 in contrast to a male predominance in the age group 36-55. ^7,8,14,15,16^

### Risk factors

A male predominance in risk factors such as dyslipidemia, smoking, arterial hypertension, coronary heart disease, substance and alcohol abuse was demonstrated in the presented cohort. This is in line with previous studies.^2,3,7–9,16,17^ While overweight was more frequent in men, women had a higher obesity rate. While some studies report higher obesity rates in female young stroke patients^3,9,18^, the Helsinki Young Stroke Registry found no sex difference regarding obesity.^7^ Unlike prior reports^7,18,19^, we did not find sex difference in the prevalence of migraine.

The subgroup analysis in patients aged 36-55 years revealed similar risk factors compared to the overall study cohort. In contrast, in the 16-35 year age group the sexes only differed in sleep-disordered breathing, and the female sex specific risk factors. Sleep-disordered breathing was more prevalent in men. A male predominance of sleep-disordered breathing was also found in the sifap1 study^18^ and the Helsinki Young Stroke Registry, but both did not further analyze different age groups in the young.^7^

The sex specific risk factors, such as pregnancy and puerperium, were almost exclusively present in the age group 16-35. This may to some extent explain the higher incidence of stroke in women in childbearing age.^14,20,21,22^

### Stroke etiology

Unlike prior reports, we did not find sex specific differences in stroke etiology classified by TOAST criteria. Previous studies described a higher rate of large artery atherosclerosis,^7,16-17^ and small vessel disease^7,16,23^ in young men compared to young women, while stroke in women was more often classified as “other determined” or undetermined cause.^16,17,23,24^ In accordance with previous studies we found a higher rate of undetermined stroke etiologies in young stroke patients despite an extensive etiological work-up including cerebral MRI, transesophageal echocardiography, at least 3 weeks of long-term ECG, and hematological work-up.

The finding of a higher prevalence of stroke risk factors in young men likely explains that women tend to have a higher RoPE-score (> 7) than men. Future studies should therefore address whether stroke in the young is more often due to paradox embolism in women than in men. It should also be addressed, whether women benefit more from a mechanical closure of the PFO.

### Stroke severity, infarct localization, treatment, and outcome

Neither stroke severity expressed by the NIHSS, nor the affected vascular territory differed between sexes. In the absence of a significant difference in etiology, stroke severity and localization, it is not surprising that no sex-related differences in the applied treatment modalities and the three-month clinical outcome expressed with the mRS was demonstrated. Only one previous study analyzed sex differences in the localization of stroke in young patients, reporting more supratentorial strokes in women.^19^

Female sex was a predictor of worse outcome (mRS>2) at discharge in patients aged 31-50 years after a first ever AIS in a Spanish study^25^, while age and male sex were associated with other arterial events, but not with stroke in the FUTURE study.^23^

### Limitations

This study is not population-based. Only patients from the area covered by our Stroke Center were included. However, in our region most young stroke patients are admitted to our stroke center, thus decreasing the risk for a selection bias due to the tertiary care setting. An ethnic bias must also be taken into account, since >95% of the population in the surrounding of the Bernese Stroke Center is Caucasian, and hardly any Africans/Asians.

We did not consider socioeconomic factors in our study, thus limiting the analysis of sex aspects, nor did we include patient centered outcome measures such as quality of life.

Our aim was to identify potential differences using univariate comparisons without adjustment for multiple testing. We did not perform multiple testing due to the limited patients of the subgroups, especially of the youngest, and the non-population-based study design. Hence, our findings should be regarded as hypothesis generating and need to be replicated in a population-based larger study.

In order to determine further characteristics of young stroke patients, a control group of young adults without stroke would be necessary. In this respect, a preliminary comparison of our cohort group with peers in the general Swiss population was made. The characteristics of the latter are regularly assessed by national surveys. The results are attached in the supplemental material.

## Data Availability

All relevant data are within the manuscript and its Supporting Information files.

## Abbreviations and Acronyms

AIS: acute ischemic stroke
IAT: intra-arterial treatment
IVT: intravenous thrombolysis
mRS: modified Rankin Scale
NIHSS: National Institutes of Health Stroke Scale
PFO: persistent foramen ovale
RoPE: risk of paradoxical embolism
rtPA: recombinant tissue plasminogen activator
TIA: transient ischemic attack
TOAST: Trial of Org 10172 in Acute Stroke Treatment

## Acknowledgements

We thank all patients for participation.

Special thanks go to Marianne Kormann (study nurse) and all research fellows.

## Figure Titles and Legends

**Figure 1:** Y indicates years; CHD, coronary heart disease; TIA, transient ischemic attack

**Figure 2:** Y indicates years

1: Large artery atherosclerosis/macroangiopathy

2: Cardiac embolism

3: Small vessel disease/microangiopathies

4: Other determined etiology

5. Undetermined etiology, including undetermined despite complete evaluation, undetermined without complete evaluation and multiple possible etiologies

## Supplemental material

We compared the risk factor prevalence in our study population with the age matched general Swiss population based on data of the National Health Survey.^26^

As illustrated in Table 3, the overall prevalence of risk factors in our cohort was higher than in the general population,^26^ which is also consistent with previous reports.^9,27^

**Tab. 3:** Risk factors according to sex in study and general Swiss population

| **Traditional risk factors** | **<u>Study population (age 36-55y)</u>** |  | **<u>General Swiss population (age 35-54y) \*</u>** |  |
| --- | --- | --- | --- | --- |
|  | **Female n=270 (%)** | **Male n=419 (%)** | **Female (%)** | **Male (%)** |
| Arterial hypertension | 37.4 | 41.2 | 12.7 - 20.0 | 18.7 - 27.1 |
| Dyslipidemia | 57.8 | 61.3 | 8.7 - 13.7 | 11.6 - 22.0 |
| Smoking | 35.0 | 45.6 | 24.6 - 26.6 | 30.6 - 36.7 |
| Diabetes | 11.3 | 9.4 | 3.0 - 3.1 | 1.0 - 3.6 |
| Overweight | 22.8 | 45.8 | 19.3 - 24.0 | 39.9 - 42.9 |
| Obesity | 28.5 | 20.4 | 9.1 - 12.1 | 11.0 - 14.4 |

\* According to the Swiss Federal Statistical Office<sup>26</sup>
Y indicates years

When comparing the frequencies of traditional risk factors in the general Swiss population based on the Health Survey of 2017 published by the Swiss Federal Statistical Office with our study population, it should be noted that the data assessment is methodologically different.

## References

1. Katan M, Luft A. Global Burden of Stroke. Semin Neurol 2018;38:208–11.

2. Maaijwee NAMM, Rutten-Jacobs LCA, Schaapsmeerders P, Van Dijk EJ, De Leeuw F-E. Ischaemic stroke in young adults: Risk factors and long-term consequences. Nat. Rev. Neurol. 2014;10:315–25.

3. Boot E, Ekker MS, Putaala J, Kittner S, De Leeuw FE, Tuladhar AM. Ischaemic stroke in young adults: A global perspective. J. Neurol. Neurosurg. Psychiatry. 2020;91:411–7.

4. Ekker MS, Boot EM, Singhal AB, et al. Epidemiology, aetiology, and management of ischaemic stroke in young adults. Lancet Neurol 2018;17:790–801.

5. Petrea RE, Beiser AS, Seshadri S, Kelly-Hayes M, Kase CS, Wolf PA. Gender differences in stroke incidence and poststroke disability in the framingham heart study. Stroke 2009;40:1032–7.

6. Yafasova A, Fosbøl EL, Christiansen MN, et al. Time trends in incidence, comorbidity, and mortality of ischemic stroke in Denmark (1996-2016). Neurology 2020;95:e2343–53.

7. Putaala J, Metso AJ, Metso TM, et al. Analysis of 1008 consecutive patients aged 15 to 49 with first-ever ischemic stroke the Helsinki young stroke registry. Stroke 2009;40:1195–203.

8. Putaala J, Yesilot N, Waje-Andreassen U, et al. Demographic and geographic vascular risk factor differences in european young adults with ischemic stroke: The 15 cities young stroke study. Stroke 2012;43:2624–30.

9. Aigner A, Grittner U, Rolfs A, Norrving B, Siegerink B, Busch MA. Contribution of Established Stroke Risk Factors to the Burden of Stroke in Young Adults. Stroke 2017;48:1744–51.

10. Brott T, Adams HP, Olinger CP, et al. Measurements of acute cerebral infarction: A clinical examination scale. Stroke 1989;20:864–70.

11. Adams H., Adams H., Bendixen B., et al. Classification of Subtype of Acute Ischemic Stroke. Stroke 1993;23:35–41.

12. Bamford JM, Sandercock PAG, Wariow CP, Slattery J. Interobserver agreement for the assessment of handicap in stroke patients: To the editor. Stroke 1989;20:828.

13. Kent DM, Thaler DE. The Risk of Paradoxical Embolism (RoPE) Study: Developing risk models for application to ongoing randomized trials of percutaneous patent foramen ovale closure for cryptogenic stroke. Trials 2011;12:185.

14. Sultan S, Elkind MS V. The growing problem of stroke among young adults. Curr Cardiol Rep 2013;15:421.

15. Smajlovic D. Strokes in young adults: Epidemiology and prevention. Vasc Health Risk Manag 2015;11:157–64.

16. Rolfs A, Fazekas F, Grittner U, et al. Acute cerebrovascular disease in the young: The stroke in young fabry patients study. Stroke 2013;44:340–9.

17. Zhang B, Pu S, Zhang W, et al. Sex differences in risk factors, etiology, and short-term outcome of cerebral infarction in young patients. Atherosclerosis 2011;216:420–5.

18. Von Sarnowski B, Schminke U, Grittner U, et al. Posterior versus Anterior Circulation Stroke in Young Adults: A Comparative Study of Stroke Aetiologies and Risk Factors in Stroke among Young Fabry Patients (sifap1). Cerebrovasc Dis 2017;43:152–60.

19. Ji R, Schwamm LH, Pervez MA, Singhal AB. Ischemic stroke and transient ischemic attack in young adults: Risk factors, diagnostic yield, neuroimaging, and thrombolysis. Arch Neurol 2013;70:51–7.

20. Van Alebeek ME, De Heus R, Tuladhar AM, De Leeuw FE. Pregnancy and ischemic stroke: A practical guide to management. Curr Opin Neurol 2018;31:44–51.

21. Salisbury M, Pfeffer G, Yip S. Stroke in young women. Can J Neurol Sci 2011;38:404–10.

22. Del Zotto E, Giossi A, Volonghi I, Costa P, Padovani A, Pezzini A. Ischemic Stroke during Pregnancy and Puerperium. Stroke Res Treat 2011;2011:1–13.

23. Yesilot Barlas N, Putaala J, Waje-Andreassen U, et al. Etiology of first-ever ischaemic stroke in European young adults: The 15 cities young stroke study. Eur J Neurol 2013;20:1431–9.

24. Nakagawa E, Hoffmann M. Young women’s stroke etiology differs from that in young men: An analysis of 511 patients. Neurol Int 2013;5:37–40.

25. Martínez-Sánchez P, Fuentes B, Fernández-Domínguez J, et al. Young women have poorer outcomes than men after stroke. Cerebrovasc Dis 2011;31(:455–63.

26. Bundesamt für Statistik. Gesundheitsverhalten. Available from: http://www.portal-stat.admin.ch/sgb2017/files/de/02a.xml

27. Goeggel Simonetti B, Mono ML, Huynh-Do U, et al. Risk factors, aetiology and outcome of ischaemic stroke in young adults: the Swiss Young Stroke Study (SYSS). J Neurol 2015;262:2025–32

